# Digital language markers distinguish frontal from right anterior temporal lobe atrophy in frontotemporal dementia

**DOI:** 10.1101/2024.08.29.24312807

**Authors:** Jet M.J. Vonk, Brittany T. Morin, Janhavi Pillai, David Rosado Rolon, Rian Bogley, David Paul Baquirin, Zoe Ezzes, Boon Lead Tee, Jessica DeLeon, Lisa Wauters, Sladjana Lukic, Maxime Montembeault, Kyan Younes, Zachary Miller, Adolfo M. García, Maria Luisa Mandelli, Virginia E. Sturm, Bruce L. Miller, Maria Luisa Gorno-Tempini

## Abstract

**Background and Objectives:** Within frontotemporal dementia (FTD), the behavioral variant (bvFTD) characterized by frontal atrophy, and semantic behavioral variant (sbvFTD) characterized by right anterior temporal lobe (rATL) atrophy, present diagnostic challenges due to overlapping symptoms and neuroanatomy. Accurate differentiation is crucial for clinical trial inclusion targeting TDP-43 proteinopathies. This study investigated whether automated speech analysis can distinguish between FTD-related rATL and frontal atrophy, potentially offering a non-invasive diagnostic tool.

**Methods:** In a cross-sectional design, we included 40 participants with FTD-related predominant frontal atrophy (n=16) or predominant rATL atrophy (n=24) and 22 healthy controls from the UCSF Memory and Aging Center. Using stepwise logistic regression and receiver operating characteristic (ROC) curve analysis, we analyzed 16 linguistic and acoustic features that were extracted automatically from audio-recorded picture description tasks. Neuroimaging data were analyzed using voxel-based morphometry to examine brain-behavior relationships of regional atrophy with the features selected in the regression models.

**Results:** Logistic regression identified three features (content units, lexical frequency, familiarity) differentiating the overall FTD group from controls (AUC=.973), adjusted for age. Within the FTD group, five features (adpositions/total words ratio, arousal, syllable pause duration, restarts, words containing ‘thing’) differentiated frontal from rATL atrophy (AUC=.943). Neuroimaging analyses showed that semantic features (lexical frequency, content units, ‘thing’ words) were linked to bilateral inferior temporal lobe structures, speech and lexical features (syllable pause duration, adpositions/total words ratio) to bilateral inferior frontal gyri, and socio-emotional features (arousal) to areas known to mediate social cognition including the right insula and bilateral anterior temporal structures. As a composite score, this set of five features was uniquely associated with rATL atrophy.

**Discussion:** Automated speech analysis effectively distinguished the overall FTD group from controls and differentiated between frontal and rATL atrophy. The neuroimaging findings for individual features highlight the neural basis of language impairments in these FTD variants, and when considered together, underscore the importance of utilizing features’ combined power to identify impaired language patterns. Automated speech analysis could enhance early diagnosis and monitoring of FTD, offering a scalable, non-invasive alternative to traditional methods, particularly in resource-limited settings. Further research should aim to integrate automated speech analysis into multi-modal diagnostic frameworks.

## Introduction

Frontotemporal dementia-spectrum disorders (FTD) encompass various neurodegenerative clinical syndromes, each associated with varying probabilities of a spectrum of pathological changes called frontotemporal lobar degeneration. Behavioral variant FTD (bvFTD) is the most prevalent FTD syndrome,^1^ clinically characterized by behavioral and personality changes, and neuroanatomically by predominant atrophy in the frontal lobes, generally right more than left-lateralized.^2,3^ Within bvFTD, anterior temporal lobe (ATL) atrophy is variable and, if present, is typically linked to Pick’s pathology when in conjunction with frontal atrophy, or TDP-43 proteinopathy type C (TDP-C) pathology when atrophy is isolated to the right ATL (rATL).^4,5^ Since isolated rATL atrophy is highly predictive of TDP-C pathology, recent studies have highlighted the importance of characterizing and isolating this syndrome from other FTD syndromes.^6^ To this end, new criteria have been proposed for a syndrome named semantic behavioral variant FTD (sbvFTD), characterized by rATL atrophy and non-verbal semantic memory impairment related to socioemotional concepts.^3,7–9^ Differential diagnosis of sbvFTD remains challenging due to the clinical overlap in socio-emotional functioning with bvFTD, and lexical semantic deficits with the left-lateralized semantic variant of primary progressive aphasia (svPPA).^3,7,10^ While distinguishing rATL and svPPA-related left ATL (lATL) atrophy is theoretically interesting, it is less clinically crucial since both syndromes (together classified as semantic dementia) share common pathology. However, accurately differentiating bvFTD and sbvFTD is clinically relevant for selecting participants for trials targeting TDP-43 proteinopathies as predominantly rATL atrophy is specifically linked to TDP-C, while bvFTD-associated frontal atrophy varies in neuropathology.^11^

In the current bvFTD criteria, clinical changes manifest as a wide range of symptoms, including loss of empathy and ability to form social relationships, disinhibition and impulsive behavior, loss of interest in activities and hobbies, decreased motivation and initiative, changes in food preferences and eating habits, apathy and lack of emotion, and/or decline in personal hygiene and grooming.^4,12^ SbvFTD with isolated rATL atrophy is instead described as having a more circumscribed set of symptoms related to loss of understanding of socioemotionally relevant, mainly non-verbal semantic concepts, such as person-specific biographical, facial expression, and emotion knowledge^3,13^ This type of semantic impairment most often results in loss of understanding of people’s identity and emotions, manifesting clinically as loss of empathy and difficulties recognizing familiar people. Difficulty interpreting non-verbal concepts (e.g., food and taste-related semantics) and extralinguistic social cues (e.g., sarcasm), repetitive behaviors, apathy, and verbal semantic deficits can be present, likely in relation to compromised functional connectivity or atrophy in orbitofrontal regions (linked to behavioral changes) and lATL regions (linked to semantic changes).^3,7,10^

The proposed sbvFTD criteria are in the early stages of development and serve as a starting point to highlight the need for a better understanding of the clinical manifestations of rATL damage, as well as its differentiation from behavioral symptoms caused by mainly frontal atrophy.^3,7^ There is a pressing need for new tools that can distinguish between symptoms of frontal atrophy associated with bvFTD and symptoms of predominantly rATL atrophy associated with sbvFTD. While both bvFTD and sbvFTD share behavioral symptoms, sbvFTD uniquely falls on the semantic dementia spectrum, introducing distinct linguistic and socio-emotional impairments that could facilitate clearer differentiation between bvFTD and sbvFTD.

Automated connected speech analysis using natural language processing (NLP) offers promising avenues for identifying early cognitive markers of dementia.^14^ This method involves automatically extracting various linguistic and acoustic features from audio recordings and transcriptions (e.g., responses to a picture description task or a free prompt). Previous connected speech research has largely focused on Alzheimer’s disease, mild cognitive impairment, and PPA, with less studies focused on behavioral FTD variants.^15–17^ Notably, one prior study demonstrated the potential of automated connected speech analysis in distinguishing between right and left ATL atrophy.^18^

The present study focused on investigating the clinically-relevant question of whether automated speech analysis can effectively differentiate between rATL atrophy and frontal atrophy in FTD. Language is a multifaceted communication system composed of hundreds of interrelated linguistic and acoustic features that work together to convey meaning, structure, tone, emotion, prosody, pragmatics, and speaker intent. The interplay of these elements not only provides the structural and functional framework of spoken language but also encodes social and contextual information, speaker identity, and cognitive states. For this study, we investigated linguistic and acoustic features based on clinical observations of speech and language in bvFTD, PPA, and semantic dementia.^18–20^ In particular, we focused on 21 features that may mark elements in speech that capture 1. behavior (e.g., loss of interest, decreased initiative): number of t-units (i.e., main clause plus any subordinate clauses that may be attached to it), average t-unit length, number of verb phrases per t-unit; 2. socio-emotional aspects: valence, arousal; 3. semantic deficits (e.g., word-finding difficulties, coherence, and topic deviation): words containing ‘thing’/total words ratio, age of acquisition, familiarity, ambiguity, concreteness, lexical frequency, number of content units, informativeness ratio, pause rate, number of silent pauses, average syllable pause duration, number of restarts; or 4. less complex language use: adpositions/total words ratio, noun/verb ratio, content/function word ratio, number of content words. Our hypotheses about brain-behavior relationships were that behavioral features would link to bilateral frontal brain regions, socio-emotional features to the rATL, and semantic features to the lATL. We did not hypothesize specific brain regions for features indicative of less complex language use, beyond anticipating associations with atrophy in the frontal and/or temporal lobes.

## Methods

### Participants

Participants were part of the University of California, San Francisco (UCSF) Memory and Aging Center (MAC) database. Study sample selection for rATL and frontal atrophy patterns followed procedures as described by Younes et al.^3^ In short, we identified all individuals meeting a clinical diagnosis of sbvFTD, bvFTD and/or svPPA based on Neary-FTD, Neary-Semantic, bvFTD, svPPA, and the newly established sbvFTD diagnostic criteria,^6,12,21,22^ who had research visits between 1998 and 2023 in the UCSF MAC database. In addition to the exclusion criteria in Younes et al., individuals were excluded in this study if they did not complete a picture description task to elicit connected speech, were not native speakers of English, and did not have a brain MRI scan within one year of their cognitive evaluation. Based on anatomical criteria, individuals were included if they had peak atrophy in either the frontal lobe or rATL, and if the ratio of temporal to frontal atrophy showed more atrophy in their peak atrophy lobe compared to the contrasting lobe. Additionally, a group of cognitively healthy control participants was selected from the MAC database. Figure 1 shows a flowchart of participant selection with a final sample of 62 participants. All participants or caregivers provided informed consent following procedures aligned with the Declaration of Helsinki, and the study was approved by the UCSF Institutional Review Board.

**Figure 1.**
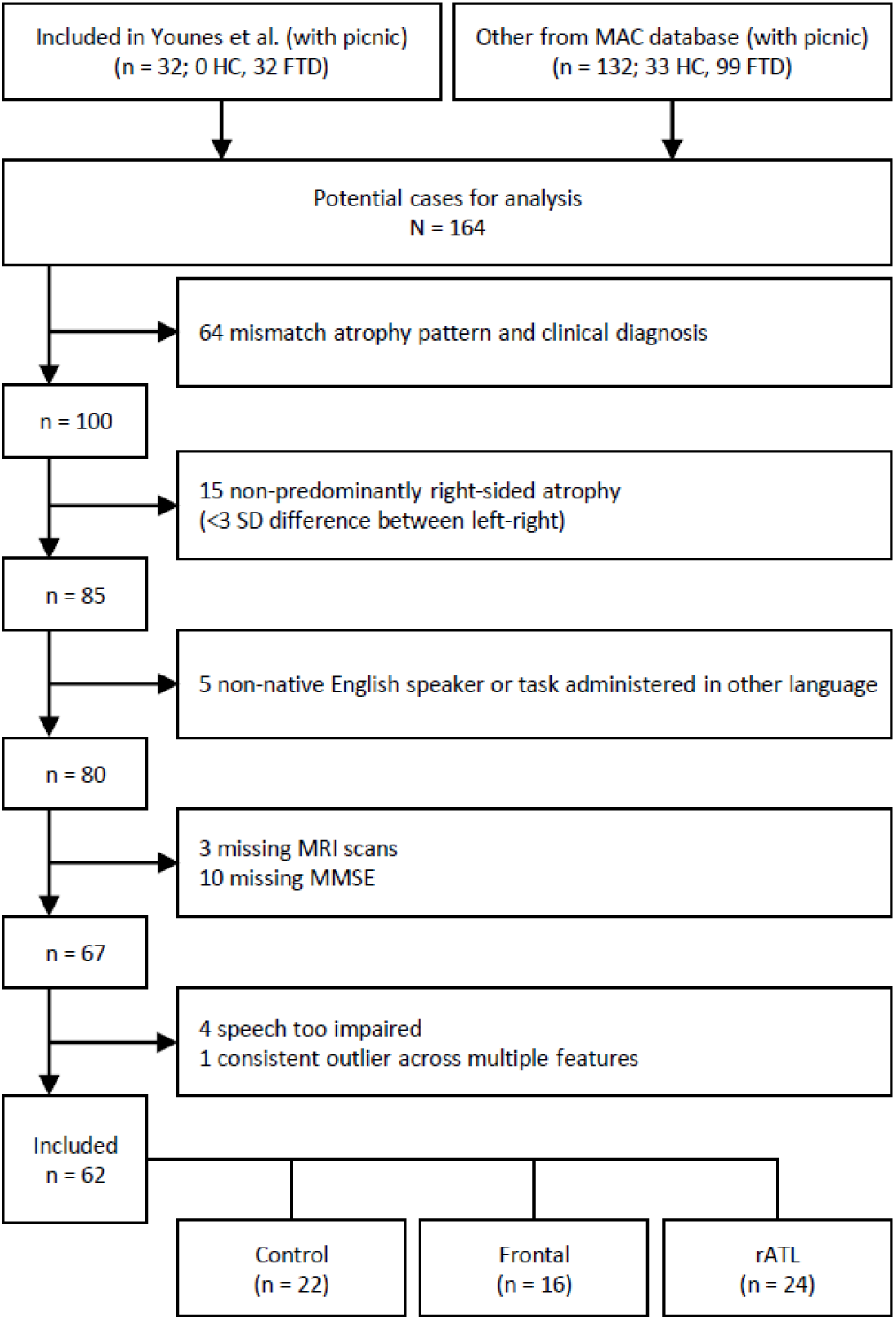
Participant selection flowchart.

### Speech Task and Automated Speech and Language Analysis

Speech samples were collected using the ‘picnic scene’ picture description task of the Western Aphasia Battery.^23^ Participants were tested individually in a quiet room at a table with the investigator seated opposite of them and their speech was recorded. Participants were instructed to: “Tell me what you see. Talk in sentences.” Participants were allowed approximately three minutes to complete the task; in our sample, participants completed the picnic picture description task within 1-2 minutes with an average time of 1m20s.

We developed an in-house pipeline for automated connected speech transcription and language analysis, named the Clinical Linguistic Automated Speech Pipeline (CLASP). Digital audio recordings of participants’ picnic descriptions were converted uniformly to .wav files (henceforth: acoustic files). Acoustic files were manually diarized by editing out any speech not belonging to the participant using Audacity version 3.5.1 (author BTM). Subsequently, acoustic files were denoised using Adobe Premiere Pro DeNoise at 50% (i.e., the amount of processing applied to the audio signal). In the next step, acoustic files were automatically transcribed using OpenAI’s large Whisper model and manually checked for quality in terms of correct wording, punctuation, and grammar (author BTM). Next, the pipeline utilized various NLP and computational tools to extract acoustic and linguistic features from both the audio recordings and their transcriptions; these tools include spaCy,^24^ PRAAT (via MyProsody^25^), PyDub, TAASSC,^26^ and custom scripts (see Table 1).

**Table 1.** Details of the linguistic and acoustic features extracted for speech and language analysis.

| *Feature* | *Description* | *Interpretation* | *Extraction Tool* |
| --- | --- | --- | --- |
| **Pause rate** | Ratio of the number of silent pauses (threshold >750ms) to the speech sample's total duration | Reflecting the relative frequency with which the speaker takes a long pause indicating difficulties with word retrieval and sentence planning<sup>44</sup> | Adapted code from the Python library MyProsody<sup>25</sup> |
| **Average syllable pause duration** | The mean length of filled pauses ("uhm"s), i.e., long stable syllables pronounced at a low pitch, measured in milliseconds | Reflecting interruptions in speech flow; longer filled pauses indicate more disfluency in speech | Adapted code from the Python library MyProsody<sup>25</sup> |
| **Number of restarts** | The instances where a speaker starts a word or phrase, stops, and then restarts, often to repair the sentence or choose a different word | Reflecting hesitations or corrections; more restarts indicate more disfluency in speech | Custom script based on Transcription Guidelines of the Linguistic Data Consortium<sup>45</sup> |
| **Noun/verb ratio** | The ratio of nouns to verbs used | Reflecting the richness and complexity of language use; a lower ratio indicates difficulty with retrieving nouns and a stronger reliance on verbs in simpler sentence structures | Parts-of-speech extracted using the Python library spaCy;<sup>24</sup> ratios calculated with custom script |
| **Adpositions/total words ratio** | The ratio of adpositions (prepositions and postpositions) to the total number of words | Reflecting the use of grammatical structures that show relationships between words; a lower ratio indicates a shift toward simpler, less complex language | Parts-of-speech extracted using the Python library spaCy;<sup>24</sup> ratios calculated with custom script |
| **Content/function word ratio** | The ratio of content words (nouns, verbs, adjectives, adverbs) to function words (pronouns, prepositions, conjunctions, auxiliary verbs) | Reflecting the balance between meaning-carrying words and grammatical words; a lower ratio suggests a reliance on function words that carry less semantic weight | Parts-of-speech extracted using the Python library spaCy;<sup>24</sup> ratios calculated with custom script |
| **Thing words ratio** | The ratio of non-specific 'thing' words (i.e., thing, something, anything) to the total number of words | Reflecting word-finding difficulties; a higher ratio suggests issues with semantic memory to access specific knowledge about objects, people, and events, leading to reliance on non-specific terms | Custom script based on Clarke et al. (2021)<sup>17</sup> |
| **Age of acquisition** | The mean age at which the words used are typically learned | Reflecting lexical richness; the use of words with an earlier age of acquisition indicates a preference for simpler, more easily accessible vocabulary | Custom script based on linguistic database by Kuperman et al. (2012)<sup>46</sup> |
| **Familiarity** | The average rating of how familiar the words are to the general population | Reflecting lexical richness; the use of words with higher familiarity indicates a preference for simpler, more easily accessible vocabulary | Custom script based on linguistic database by Brysbaert et al. (2019)<sup>47</sup> |
| **Lexical frequency** | The mean lexical frequency of the words used, i.e., how often these words are generally used in daily language | Reflecting lexical richness; the use of words with higher lexical frequency indicates a preference for simpler, more easily accessible vocabulary | Custom script based on linguistic database by Brysbaert et al. (2009)<sup>48</sup> |
| **Arousal** | The average level of emotional intensity of the words used | Reflecting the overall level of emotional intensity of the words used; a deviation of neutral arousal in a description task indicates emotional alterations | Custom script based on linguistic database by Warriner et al. (2013)<sup>49</sup> |
| **Valence** | The mean emotional value (positive or negative) of the words used | Reflecting the overall sentiment of the words used; a deviation of neutral valence in a description task indicates emotional alterations | Custom script based on linguistic database by Warriner et al. (2013)<sup>49</sup> |
| **Number of T-units** | The total number of T-units (shortest grammatically allowable sentence), i.e., a main clause and any subordinate clause(s) | Reflecting syntactic complexity; a lower number of T-units indicate an overall decrease in speech output | Tool for the Automatic Analysis of Syntactic Sophistication and Complexity (TAASSC 1.3.8)<sup>26</sup> |
| **Average T-unit length** | The mean length of T-units in terms of words | Reflecting syntactic complexity; shorter T-units indicate simpler sentences | Tool for the Automatic Analysis of Syntactic Sophistication and Complexity (TAASSC 1.3.8)<sup>26</sup> |
| **Number of content units** | The total count of expected key information units for the WAB Picnic Description Task | Reflecting the completeness and informativeness of the picture description; a lower number of content units indicates a less complete description | Custom script based on Josephy-Hernandez et al. (2023)<sup>50</sup> |
| **Informativeness ratio** | The ratio of relevant content units to the total number of words | Reflecting the efficiency and focus of the description in conveying important information; a lower ratio indicates less efficiency | Custom script based on Josephy-Hernandez et al. (2023)<sup>50</sup> |

### Statistical Analysis

Sample characteristics and feature distributions were analyzed with descriptive statistics, general linear models, chi-square tests, and Pearson’s correlation coefficients. The distributions of each feature were examined for outliers by each diagnostic group (defined as values ≥2.5 SDs from the feature’s group mean) for measurement error or data entry error; we did not remove individual outlier values. We closely examined individuals whose data included outliers on multiple features and removed one participant who was an outlier on four features (all other participants had outliers on at most two features). We also excluded four individuals who were too impaired to complete the task, producing fewer than 25 words (see flowchart Figure 1). We applied a logarithmic transformation to 10 features to render skewed distributions more normal and diminish effects of outliers. To assess the linearity assumption of continuous predictors in a logistic regression model, we ran Box-Tidwell tests; all variables passed these tests for both outcomes (controls vs. FTD, or frontal vs. rATL atrophy). There were no missing data on the linguistic and acoustic features.

We examined multicollinearity among the 21 features (see Introduction) with a Pearson correlation table; based on a high correlation of r > |0.7| we removed five features from further consideration, including number of verb phrases per t-unit, number of content words, number of silent pauses, ambiguity, and concreteness. Thus, we included 16 linguistic and acoustic features extracted with our in-house pipeline to measure various aspects of participants’ speech and language use (Table 1).

We performed stepwise logistic regression to automatically select a reduced number of linguistic and acoustic features for building the best-performing classification model per binary outcome. Both models included age as a covariate, and the model of frontal vs. rATL atrophy additionally covaried for MMSE score as a proxy of disease severity.^27^ Sex and education were not included as covariates because there were no significant group differences. The predicted probabilities of the optimal set of features were saved as a new variable and entered into a receiver operating characteristic (ROC) curve analysis to evaluate the area under the curve (AUC). We combined the feature values of the optimal set to distinguish frontal and rATL atrophy into a composite score using z-scores, and tested this composite score in an multiple linear regression model adjusted for age and MMSE score to understand how FTD group membership (frontal vs rATL atrophy; independent variable) is associated with changes in the composite score of features (dependent variable). To put the classification performance of the set of linguistic and acoustic features into context, we also performed an ROC-AUC analysis on the Boston Naming Test,^28^ an established neuropsychological task to detect lexical-semantic difficulties.

All statistical analyses were performed in SPSS Version 28.0.1.0. Tables and figures for these analyses were generated using R Version 4.3.0 with packages: dplyr, ggplot2, furniture, summarytools, pROC, Hmisc, and corrplot. All analysis code is available at https://github.com/jmjvonk.

### Neuroimaging Analysis

We performed a Voxel-Based Morphometric (VBM) analysis using the Computational Anatomy Toolbox 12 (dbm.neuro.uni-jena.de/cat) in Statistical Parametric Mapping 12 software (fil.ion.ucl.ac.uk/spm/software/spm12) through MATLAB (version 9.14.0.2239454). MRI acquisition T1 images were acquired for all subjects with sequences, previously described, on either 1.5T (n=6),^29^ 3T (n=50),^30^ or 4T (n=2).^31^ After a visual quality check, the structural images underwent enhancement using a spatial adaptive non-local means denoising algorithm followed by bias field correction, affine transformation for alignment, and processing using SPM’s “unified segmentation” protocol. The images were then (i) segmented into gray matter, white matter, and cerebrospinal fluid and (ii) spatially normalized to the Montreal Neurological Institute (MNI) reference space via an advanced geodesic shooting technique, (iii) adjusted by the Jacobian determinants of the deformation field during spatial normalization to maintain the tissue volume integrity, and (iv) output with a uniform isotropic voxel resolution of 1.5 × 1.5 × 1.5 mm³, (v) spatially smoothed with an 8 mm full-width at half-maximum isotropic Gaussian kernel to compensate for residual anatomical variability.

We examined brain-behavior relationships between linguistic features and gray matter volume loss (atrophy). The search volume included regions of interest based on known distribution of atrophy in FTD variants using an explicit mask created with WFU PickAtlas (https://www.nitrc.org/projects/wfu_pickatlas/), including the frontal lobe, temporal lobe, anterior cingulate, frontal-temporal space, precentral gyrus, paracentral lobule, transverse temporal gyrus, parahippocampal gyrus. Using general linear models (GLM) across the whole sample (i.e., independent of diagnostic group), we examined the relationship between gray matter volume and each linguistic feature that was selected in the forward stepwise logistic regression models. Additionally, we tested the relationship between gray matter volume and the composite score of the optimal set of features that behaviorally distinguished the rATL and frontal atrophy group. All models were adjusted for age, sex/gender, scanner (3T vs non-3T), and total intracranial volume. For each computed T-contrast, the corresponding statistical map was evaluated at a peak-level uncorrected threshold of p<0.001 and a cluster-level threshold of p<0.05 family-wise error (FWE) corrected for clusters with a minimum size of 100 voxels. Visualization of peak locations of brain-behavior associations between features and atrophy was performed using MRIcroGL (https://www.nitrc.org/projects/mricrogl).

## Results

### Participants

The final selection of participants for the behavioral analyses included 16 with predominant frontal atrophy, 24 individuals with predominant rATL atrophy, and 22 control participants (n=62; see flowchart Figure 1). Participant characteristics are described in Table 2; the three groups did not differ in mean years of education nor their distribution of sex/gender, race/ethnicity, or handedness. The frontal and rATL atrophy groups did not differ in mean age, while the control group was on average slightly older. Four participants’ MRI scans, two control and two rATL, were excluded for neuroimaging analyses due to poor image quality (i.e., n=58 for neuroimaging analyses).

**Table 2.** Participant Characteristics.

| Demographic measures | Controls<br>n = 22 | Frontal atrophy<br>n = 16 | rATL atrophy<br>n = 24 | p-value |
| --- | --- | --- | --- | --- |
| Age | 72.82 (7.47, 53-88) | 64.38 (10.01, 48-77) | 66.83 (8.7, 47-87) | 0.010 |
| Sex/gender (men) | 9 (40.9%) | 6 (37.5%) | 14 (58.3%) | 0.342 |
| Years of education | 17.45 (2.09, 12-20) | 15.69 (2.87, 9-20) | 15.92 (2.7, 12-21) | 0.060 |
| Race |  |  |  | 0.627 |
| Black/African American | 1 (4.5%) | 0 (0%) | 0 (0%) |  |
| Hispanic | 0 (0%) | 0 (0%) | 0 (0%) |  |
| Mixed | 1 (4.5%) | 0 (0%) | 1 (4.2%) |  |
| Non-Hispanic White | 20 (90.9%) | 16 (100%) | 23 (95.8%) |  |
| Handedness |  |  |  | 0.167 |
| Ambidextrous | 0 (0%) | 0 (0%) | 2 (8.3%) |  |
| Left | 4 (18.2%) | 0 (0%) | 3 (12.5%) |  |
| Right | 18 (81.8%) | 16 (100%) | 19 (79.2%) |  |
| MMSE | 29.45 (0.67, 28-30) | 23 (5.73, 8-29) | 25.62 (4.34, 12-30) | <.001 |
| CDR Box Score | 0.03 (0.11, 0-0.5) | 3.78 (2.78, 0-9) | 4.5 (2.55, 1.5-10) | <.001 |
| Boston Naming Test (15-item) | 14.76 (0.56, 13-15) | 10.44 (3.98, 3-15) | 8 (3.58, 2-15) | <.001 |
| CVLT Short Delay | 12.15 (2.98, 6-16) | 4.33 (2.61, 0-9) | 4.75 (2.17, 0-8) | <.001 |
| CVLT Long Delay | 11.45 (3.35, 5-15) | 3 (3.32, 0-9) | 3.25 (2.64, 0-8) | <.001 |

**Note.** MMSE = Mini-Mental State Examination; CDR = Clinical Dementia Rating; CVLT = California Verbal Learning Test

### Automated Speech and Language Analysis

From the list of 16 linguistic and acoustic features (Table 1), the forward stepwise logistic regression model selected an optimal set of three features to classify controls versus individuals with FTD: content units, lexical frequency, and familiarity. The model demonstrated a good fit as indicated by the Omnibus test (p<.001) and explained a large proportion of the variance with a Nagelkerke R² of .803. The model achieved an accuracy of 90.3% (i.e., number of correct predictions/total number of predictions). The predicted probabilities from the logistic regression were used to generate a ROC curve with an AUC of .969 for the linguistic features alone (Figure 2A). When age was included as a covariate, the AUC increased to .973.

**Figure 2.**
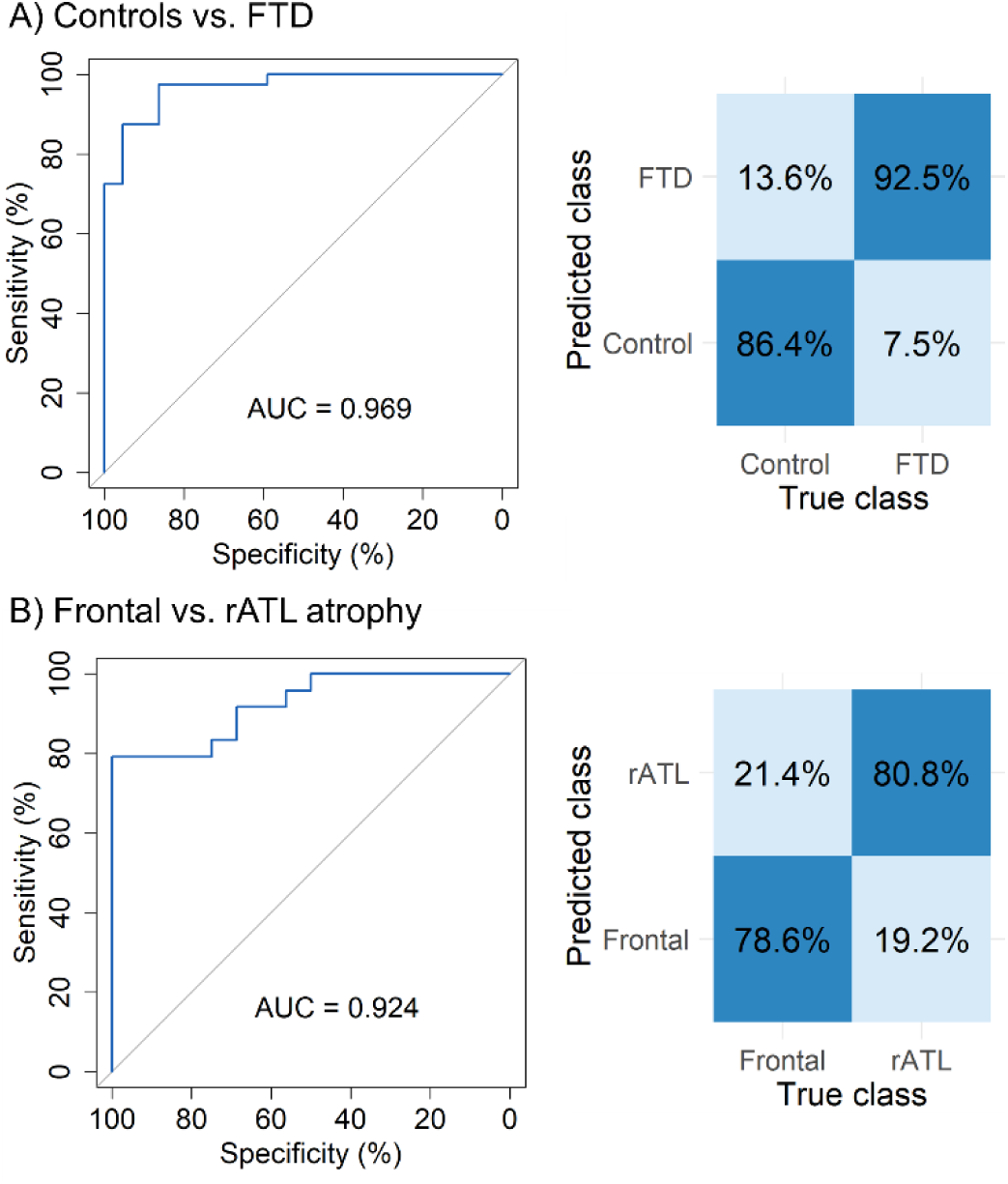
ROC curves and confusion matrices.

The forward stepwise logistic regression model to distinguish between frontal and rATL atrophy selected from the list of 16 features an optimal set of five features: adpositions/total words ratio, arousal, syllable pause duration, restarts, and words containing ‘thing’ (henceforth: “thing words”). The model demonstrated a good fit (Omnibus test p<.001), explained a substantial proportion of the variance (Nagelkerke R² = .708), and achieved an accuracy of 85.0%. The AUC was .924 for the linguistic features alone (Figure 2B). When age and MMSE scores were included as covariates, the AUC increased to .943. In comparison, an unadjusted model of the Boston Naming Test showed a moderate fit (Omnibus test p=.047), explained a small proportion of the variance (Nagelkerke R² = .130), and achieved an accuracy of 74.4% and AUC=.688. When adjusted for age and MMSE scores, the AUC improved to .813.

We created a composite score from this optimal set of five features that distinguish between frontal and rATL atrophy: adpositions/total words ratio, arousal, syllable pause duration, restarts, and thing words. Within the 40 participants with FTD, this composite score ranged from -.59 to 1.07 (m=.087, SD=.388). Multiple linear regression showed that the unstandardized coefficient B was 0.557 (SE=.97) units higher in the rATL group than the frontal group, holding covariates constant (standardized Beta = .712, p<.001; indicating a large effect size). Boxplots of performance on the selected features and the composite score per diagnostic group are shown in Figure 3.

**Figure 3.**
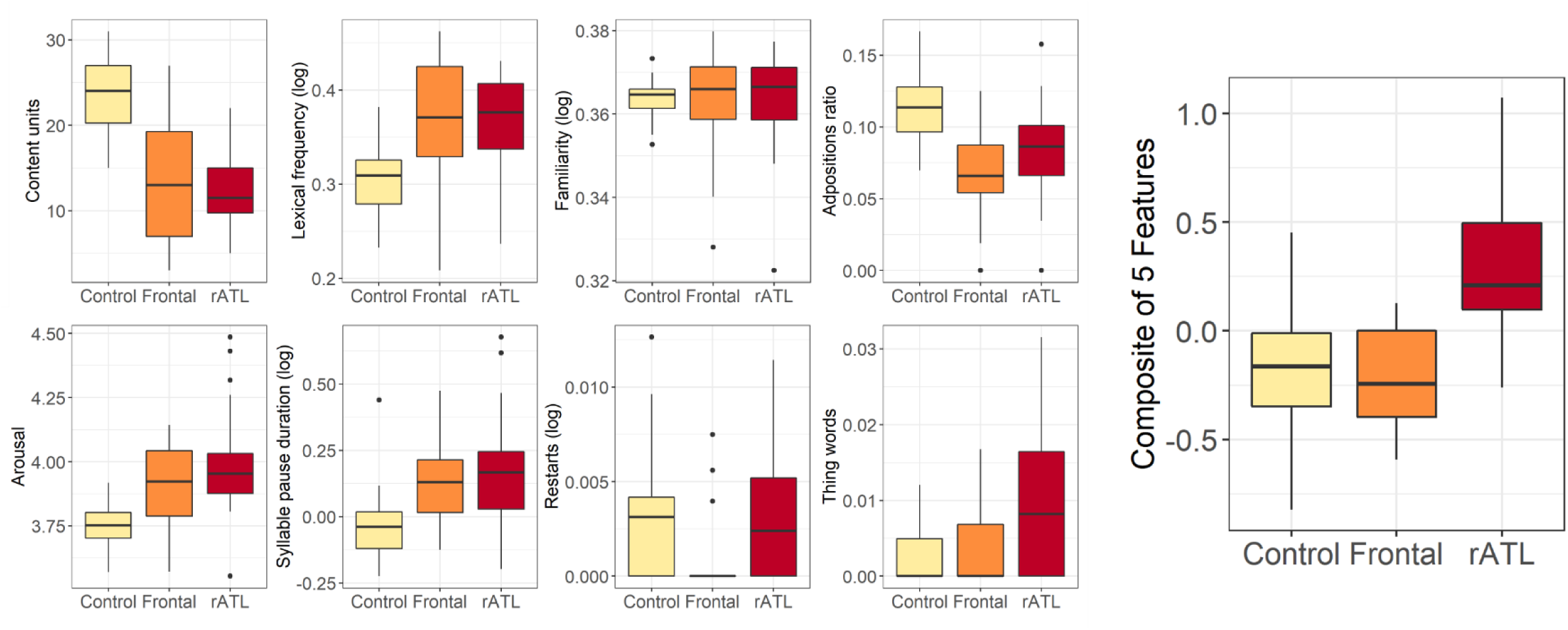
Performance on selected speech and language features across diagnostic groups; the composite score includes the optimal set of five features to distinguish between frontal and rATL atrophy: adpositions/total words ratio, arousal, syllable pause duration, restarts, and thing words.

### Brain-Behavior Associations

Table 3 displays the VBM results for the neural correlates of the speech and language features selected in the logistic regression models, and Figure 4 provides a visual representation. Across all participants, the adpositions/total word ratio was positively associated with cortical atrophy in the right orbitofrontal cortex and left inferior frontal and precentral gyri. Arousal was negatively associated with atrophy in the right insula, middle temporal gyrus, and anterior cingulate, as well as left inferior temporal structures. Content units was positively associated with atrophy in left inferior temporal structures as well as a right-sided cluster that stretched from the amygdala to the orbitofrontal cortex. Lexical frequency was negatively associated with the left inferior temporal gyrus and left middle frontal gyrus. Syllable pause duration was negatively associated with atrophy in the left pars triangularis, and thing words was negatively associated with atrophy in the right fusiform, middle temporal, and inferior temporal gyri. We did not find associations of familiarity (a priori tested direction: negative) or number of restarts (a priori tested direction: negative) with cortical atrophy in our sample. The composite score of the optimal set of features that distinguished frontal from rATL atrophy (adpositions/total words ratio, arousal, syllable pause duration, restarts, and thing words) was negatively associated with focal atrophy in the rATL, with peaks in the right fusiform gyrus, right middle temporal gyrus, and right temporal pole.

**Figure 4.**
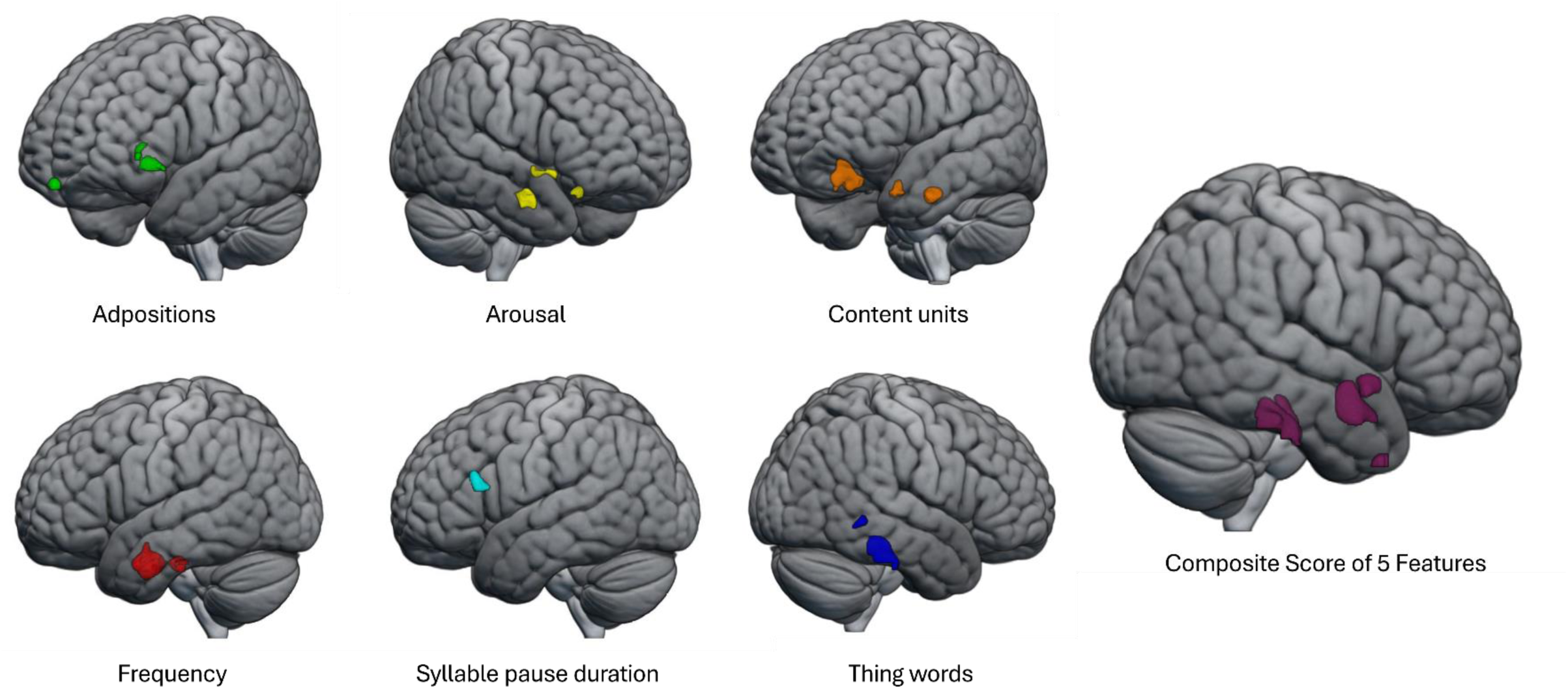
Peak locations of brain-behavior associations between features and atrophy; images are for illustration only, detailed results are summarized in Table 3.

**Table 3.**
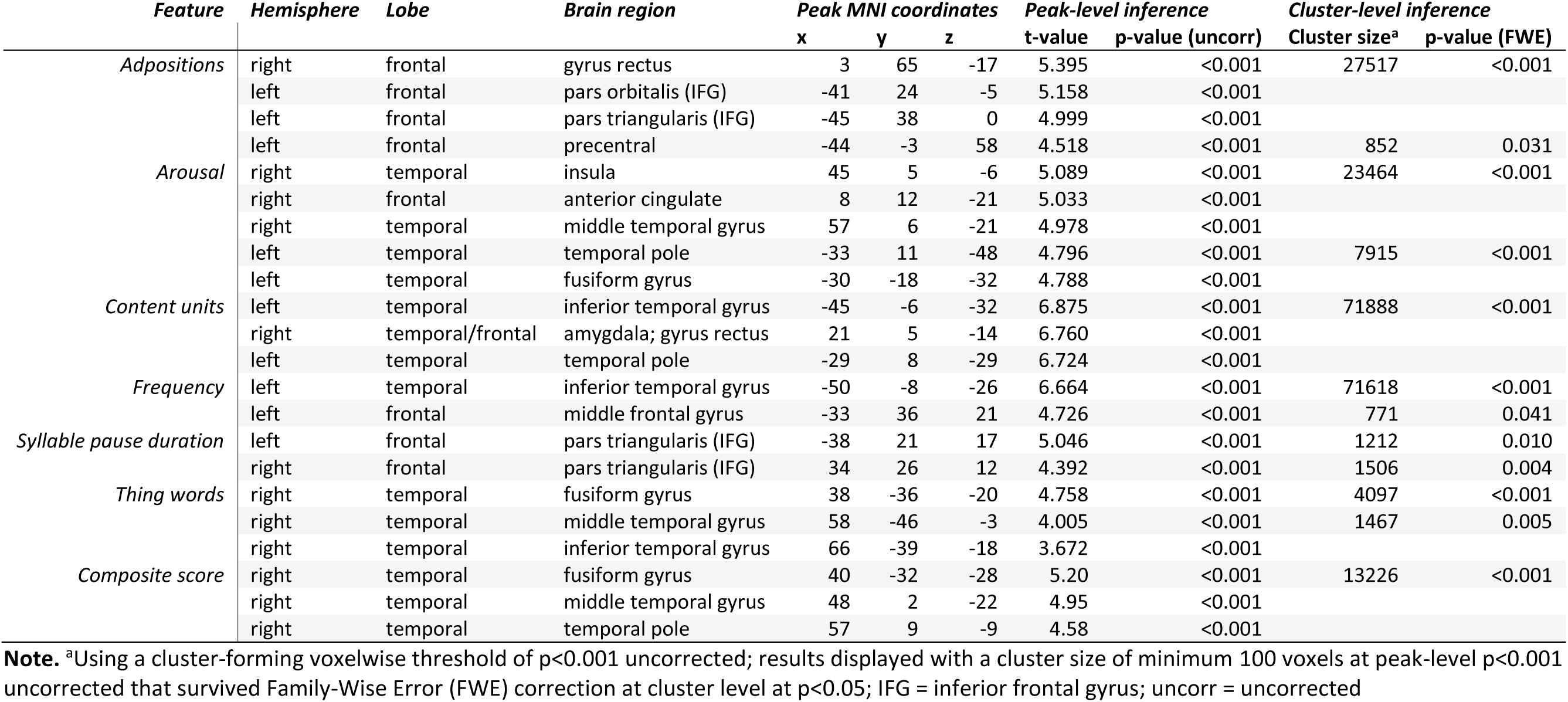
Voxel-based morphometry: neural correlates of selected speech and language features.

## Discussion

This study investigated the utility of automated speech and language analysis in differentiating between healthy controls and individuals with FTD, particularly focusing on two atrophy patterns: individuals with frontal atrophy (typically associated with bvFTD) and individuals with rATL atrophy (typically associated with sbvFTD). The results revealed a set of three linguistic and acoustic features that distinguished FTD from healthy controls, and a set of five features that discriminated frontal and rATL atrophy. This study addressed a crucial gap in the diagnostic process given the challenges in differentiating between bvFTD and sbvFTD,^3,7^ which has traditionally relied heavily on clinical judgment and neuroimaging. Moreover, the ability to distinguish controls from FTD based on speech and language features presents a promising screening tool for detecting this disease, offering quick, easily administered assessments with the potential for remote and scalable implementation outside of in-person clinical settings. By distinguishing among FTD-related frontal versus rATL atrophy, this approach could help identify patients with rATL atrophy for inclusion in molecule-targeted pharmacological clinical trials.

Previous work using automated speech analyses often has focused on comparing performance on individual linguistic and/or acoustic features, often overlooking that the true complex ‘shape’ of someone’s communication emerges from how these features interact and are tuned together in a multidimensional context. Our analytic approach using forward stepwise logistic regression centered on automatically selecting a combined set of linguistic and acoustic features for discrimination, highlighting the importance of synergistic feature combinations over isolated feature performance for predictive accuracy. Our results showed that certain combinations of speech and language features can detect the slight differences in clinical characteristics associated with frontal vs. rATL atrophy. Moreover, this set of features outperformed the Boston Naming Test, an established neuropsychological task to detect lexical-semantic difficulties. As a composite score, this selected set of features was uniquely associated with rATL atrophy. The individual features that contributed to this composite score were consistent with expected patterns of language impairment in the rATL group. For example, the rATL atrophy group used words with on average higher arousal ratings compared to the frontal atrophy and control groups. Previous work showed that atrophy in left-lateralized emotion-relevant systems in the temporal lobe relates to enhanced positive emotions in FTD,^32^ corresponding to the asymmetrical yet bilateral temporal atrophy in rATL. Moreover, the increased syllable pause duration and increased use of restarts and thing words are indicators of word-finding problems, characteristic of semantic difficulties similar to svPPA.^22^ Thus, the combined set of linguistic and acoustic features that differentiated frontal and rATL atrophy groups includes both aspects of emotional alterations and semantic impairment that together mark the sbvFTD syndrome.

The clinical presentations of bvFTD and sbvFTD have been linked to specific focal atrophy. Neuroanatomically, bvFTD is associated with atrophy particularly in the bilateral medial frontal cortex, its adjacent frontal cortex, the anterior insula, and the striatum.^2,3^ These areas are considered to play a role in social cognition, emotion alterations, motivation, and decision-making.^33,34^ SbvFTD is associated with bilateral right-more-than-left and medial-more-than-lateral neurodegeneration of the ATLs, right-more-than-left insula, right caudate, and right anterior cingulate cortex.^3,8,29^ The bilaterial ATLs have been linked extensively to their role in semantic processing;^35^ the lATL is associated with verbal semantics, while the rATL is associated with non-verbal semantics, particularly regarding visual and socioemotional knowledge.^8,36^ Our neuroimaging results show associations of linguistic and acoustic features with both left and right cortical structures, reflecting the bilateral atrophy patterns of both groups as well as the role of the left hemisphere in language and the right hemisphere in emotional processing. For example, we showed that the average arousal-value of the generated words was associated with the right insular cortex. The insula is known for its role in moderating social cognition.^37^ Moreover, the right insula in particular has been linked to moderating physiological arousal such as heart rate.^38^ Importantly, we observed that both patient groups produced on average words with a higher arousal rating than controls, in line with previous reports showing that left-lateralized atrophy in the temporal and frontal lobes in FTD can impair the ability to suppress positive emotions such as happiness.^32,37^

Lexical frequency, content units, and thing words share a common semantic focus. We found that these linguistic features were related to inferior temporal lobe structures, which are known to be a critical neural substrate for semantic processing.^39^ Notably, the association of the left inferior temporal gyrus with lexical frequency derived from automated speech analysis in an FTD population replicates the findings by Cho et al.^19^ Average syllable pause duration, which detects filled pauses (“uhm”), was associated with the pars triangularis of the inferior frontal gyrus. This region that is part of Broca’s area is considered to be involved in the production of speech, including the planning and articulation of verbal expressions. For example, a connected speech analysis by Wilson et al.^40^ showed that this region was related to speech rate. As such, the association of this region with average syllable pause duration—an acoustic feature that measures long stable syllables pronounced at a low pitch as an indicator of disfluency in speech—is theoretically supported. These findings underscore the importance of both temporal and frontal regions in different aspects of speech production and semantic processing, highlighting the neural basis of language impairments in FTD.

Our findings show that digital language markers from connected speech can classify FTD and differentiate between FTD variants, supporting the growing evidence of language and speech impairments as early indicators of neurological decline.^14^ While the majority of studies on connected speech in dementia have focused on Alzheimer’s disease,^15,17^ this study further expanded this field to FTD,^19,20,41^ showcasing the potential of connected speech analysis in differentiating between its non-language dominant subtypes and underscoring the complexity of FTD as a spectrum disorder with diverse clinical presentations. Theoretically, this study contributes to the understanding of neurodegenerative disorders, particularly in elucidating the relationship between language dysfunction and brain atrophy patterns in FTD. Practically, the identification of specific speech and language markers could aid in selection of participants into clinical trials targeting TDP-43 proteinopathies. In addition, the use of connected speech analysis as a cognitive tool offers a non-invasive and accessible method for monitoring disease progression. This application is particularly relevant for clinical trials and longitudinal studies, where objective measures of cognitive function are needed to assess the efficacy of therapeutic interventions.

A major strength of this study lies in its innovative use of automated methods to analyze connected speech. Our approach provides a scalable and quantitative method for analyzing speech and language, in contrast to time-consuming manual transcription. While we currently include a manual quality check, future research should determine whether this step is necessary or if it can be omitted without significantly affecting classification accuracy, allowing for full automation. Another strength is the inclusion of a relatively large group of individuals with rATL atrophy given the low prevalence of this disease,^42^ due to the collection of these data across many years at the specialized UCSF MAC. However, the focus on participants from a single database may not reflect the diversity of the general population. It is important to note that this study’s participant group was specifically selected to address our experimental questions as a proof-of-concept. Further research is needed to determine whether similar results, including the AUC-ROC classifications, can be replicated in more diverse, unselected patient groups. As sbvFTD and its associated rATL atrophy pattern is a relatively rare disease, our sample sizes are moderate yet comparable to other studies on rATL atrophy.^9,43^ The moderate sample size may have made the results vulnerable to potential biases inherent in speech sample collection, such as participants’ mood or environment during recording. Lastly, brain-behavior relationships in structural MRI are inherently restricted to areas with atrophy in the included disease patterns. Therefore, our findings may not capture possible associations between linguistic features and brain regions that are not affected by atrophy in the included syndromes.

Future research should aim to replicate these findings in larger and more diverse populations and focus on external validation efforts. Additionally, future work should focus on developing these measures into an easy-to-use tool with a friendly user interface to assist healthcare providers in overcoming any barriers to utilizing these techniques. Moreover, exploring the integration of speech analysis in a multi-model approach with other biomarkers and diagnostic methods, such as cognitive, neuropathological and genetic data, could enhance the accuracy and utility of these findings. Longitudinal studies may also provide insights into the progression of FTD and the potential of speech analysis in monitoring disease progression and response to treatment. The methods and findings of this study also have broader implications for research in other neurodegenerative diseases. In addition to Alzheimer’s disease and FTD, the application of automated speech and language analysis could be extended to conditions like Parkinson’s disease and multiple sclerosis, where language and cognitive impairments are also prevalent. Early diagnosis of bvFTD and sbvFTD is critical for developing management strategies, potential treatment, and an appropriate care plan. Connected speech analysis provides a complementary assessment of cognitive function to established methods, capturing nuances of the complex interplay between language and behavior that may be missed in commonly used neuropsychological testing. The identification of unique speech and language patterns linked to atrophy patterns typical of bvFTD and sbvFTD opens new avenues in understanding these conditions. Our study showed that digital language markers from a brief speech production task are useful for detecting FTD and for differentiating underlying atrophy patterns, which are typically unknown early on. Clinically, speech analysis could be used as a non-invasive and cost-effective screening tool, which could be particularly beneficial in settings where access to neuroimaging techniques is limited. Moving forward, digital innovations elevate the potential of automated speech analysis to become a quick, non-invasive, remote, scalable, and low-cost cognitive tool to enhance FTD diagnosis.

## Data availability

The conditions of our ethics approval do not permit public archiving of anonymized study data. Data generated by the UCSF MAC are available upon request. Data requests can be submitted through the UCSF MAC Resource Request form: http://memory.ucsf.edu/resources/data. Access will be granted to named individuals in accordance with ethical procedures governing the reuse of sensitive data. All requests will undergo UCSF regulated procedure thus requiring submission of a Material Transfer Agreement (MTA) which can be found at https://icd.ucsf.edu/material-transfer-and-data-agreements. No commercial use would be approved.

## Acknowledgments

This work was supported by the National Institutes of Health (M.L. Gorno-Tempini: R01NS050915, K24DC015544; U01AG052943; B.L. Miller: P50AG023501, P01AG019724; J.M.J. Vonk: R00AG066934) and NWO/ZonMw (Veni Grant project number 09150161810017). We are grateful to our patients and healthy volunteers for participating in this research.

## Notes

### Competing Interest Statement

The authors have declared no competing interest.

### Author Declarations

All participants or caregivers provided informed consent following procedures aligned with the Declaration of Helsinki, and the study was approved by the University of California San Francisco Institutional Review Board.

